# Response to the coronavirus disease 2019 (COVID-19) pandemic at private retail pharmacies in Kenya: a mixed methods study

**DOI:** 10.1101/2021.10.22.21265188

**Authors:** Peter Mugo, Audrey Mumbi, Daniella Munene, Jacinta Nzinga, Sassy Molyneux, Edwine Barasa, on behalf of PSK CRT

## Abstract

**Background:** Private retail pharmacies in developing countries present a unique channel for COVID-19 prevention. We assessed the response to the COVID-19 pandemic by pharmacies in Kenya, aiming to identify strategies for maximising their contribution to the national response.

**Methods:** We conducted a prospective mixed-methods study, consisting of a questionnaire survey (n=195), a simulated client survey (n=103), and in-depth interviews (n=18). Data collection started approximately seven months after the pandemic reached Kenya. Quantitative data were summarized using measures of central tendency and multivariable modelling done using logistic regression. Qualitative analysis followed a thematic approach.

**Results:** The initial weeks of the pandemic were characterized by fear and panic among service providers and a surge in client flow. Subsequently, 61% of pharmacies experienced a dip in demand to below pre-pandemic levels and 31% reported challenges with unavailability, high price, and poor-quality of products. Almost all pharmacies were actively providing preventive materials and therapies; educating clients on prevention measures; counselling anxious clients; and handling and referring suspect cases. Fifty-nine pharmacies (55% [95% CI 45-65%]) reported ever receiving a client asking for COVID-19 testing and a similar proportion supported pharmacy-based testing. For treatment, most pharmacies (71%) recommended alternative therapies and nutritional supplements such as vitamin C; only 27% recommended conventional therapies such as antibiotics. While 48% had at least one staff member trained on COVID-19, a general feeling of disconnection from the national program prevailed.

**Conclusions:** Private pharmacies in Kenya were actively contributing to the COVID-19 response, but more deliberate engagement, support and linkages are required. Notably, there is an urgent need to develop guidelines for pharmacy-based COVID-19 testing, a service that is clearly needed and which could greatly increase test coverage. Roll-out of this and other pharmacy-based COVID-19 programs should be accompanied with implementation research in order to inform current and future pandemic responses.

## Introduction

The coronavirus disease 2019 (COVID-19) pandemic is an ongoing global health emergency [1, 2]. As of September 2021, over 230 million cases had been confirmed globally resulting in over 4.7 million deaths [3]. For the better part of 2020, non-pharmaceutical interventions such as early case detection, contact reduction, and hygiene measures were the main prevention strategies [4, 5]. In early 2021, following unprecedented public-private collaboration and speed in research and development, a number of highly effective vaccines became available [6, 7].

Non-pharmaceutical supportive care remains the key approach to treatment and management [8-10]. Among hospitalized patients, pharmaceuticals that have been shown to be effective include corticosteroids such as dexamethasone, antivirals such as remdesivir, and immunomodulators such as tocilizumab [8, 11, 12]. In the outpatient setting, pharmaceuticals used to manage mild COVID-19 cases include those targeting fever and pain such as ibuprofen, and those aiming to relieve respiratory congestion [9][12]. Other more specific agents that have been tried, such as hydroxychloroquine, ivermectin, and azithromycin, are discouraged [9, 12]. The World Health Organization and the US National Institutes of Health recommend that such unproven therapies be used only within the context of clinical trials [13, 14].

It is predicted that follow-up waves of COVID-19 infection may continue indefinitely [15]. Strategies are therefore required to enhance and sustain coverage, cost-effectiveness, and efficiency of prevention and treatment interventions. Private retail pharmacies (also referred to as community pharmacies) are a unique channel through which COVID-19 prevention and treatment interventions can be delivered, since they are often the first or only point of contact with the healthcare system in developing countries [16-18]. The main reasons for preferential care-seeking at pharmacies, compared to health facilities, include greater accessibility, lower cost, and greater perceived privacy [19, 20]. The International Pharmaceutical Federation highlights key areas where the pharmacy sector can support the pandemic response, including: management of the medication supply chain, patient education, identification and referral of patients to other health care providers, and provision of prevention materials and therapies [21, 22]. During the COVID-19 pandemic, pharmacies in many settings, including Kenya, were categorized as essential services hence they were exempted from closure [23].

The first COVID-19 case in Kenya was reported on 13^th^ March 2020, and as of 12^th^ October 2021, there were 251,248 confirmed cases and 5,190 deaths [24]. A number of control measures have been instituted by the government at different times depending on rates of infection and mortality [23, 25, 26]. Vaccine roll-out in Kenya started in March 2021 [27]. We assessed the preparedness and response to the COVID-19 pandemic at private retail pharmacies in Kenya, aiming to identify strategies for maximising their contribution to the national response.

## Methods

### Study design

We conducted a cross-sectional descriptive study, including a questionnaire survey, a simulated client survey, and in-depth interviews. A mixed-methods approach was used in data collection and analysis: insights from interviews were used to refine the survey tools and probe quantitative data; and survey data were used to refine the interview guide and qualitative data analysis. Given the physical distancing requirements at the time, data collection was done remotely.

### Study settings and populations

The study was conducted in three counties of Kenya (Nairobi, Mombasa and Kisumu), selected purposively to represent the main urban centres. As of June 2020, the three counties together contributed 32% (1,602/5,033) of licensed pharmacies [28] and 78% (2,229/2,862) of reported COVID-19 cases [29] [24].

We targeted pharmacies that had participated in a pharmacy HIV prevention (PHP) study about a year previously (manuscript in preparation). Pharmacies had been selected randomly from the list of licensed pharmacies. Respondents were pharmacists (degree holders) and pharmaceutical technologists (diploma holders). Interview participants were purposively selected from among questionnaire respondents, targeting those that showed enthusiasm for the study through prompt and detailed responses. We aimed to increase variability of interview data by stratifying the sample by county, practice setting (urban vs. rural) and gender (male vs female).

### Data collection

Data collection started in November 2020 (approximately seven months after the pandemic reached Kenya) and was concluded in December 2020 (for the surveys) and April 2021 (for the interviews).

One structured online questionnaire, taking about 20-30 minutes, was completed by each participating pharmacy (**Supplementary file A**). Data on pharmacy characteristics were imported from the PHP study database.

Simulated clients (SCs) made unscheduled phone calls to participating pharmacies, mimicking a client seeking COVID-19 preventive therapies (**Supplementary file B**). The SCs (2 women and 2 men, aged 25-35 years) had previously been engaged in the PHP study and trained comprehensively over a 2-week period. For the current study, further training was provided over a 1-week period through the Teams^®^ digital meeting platform. An online debrief questionnaire was used to record observations (**Supplementary file C**). All calls were made on weekdays (Monday – Friday) between 8am and 5pm, over a 3-week period. Median [range] duration of calls was 4 [1-12] minutes. The majority (68%) of calls were made to a publicly available phone number, while the rest used a number in the study database as there was no public number. A few weeks after the simulated call, participating pharmacies were contacted to check if they suspected any clients visiting the pharmacies during the survey period to be “fake” (detection survey).

In-depth interviews were conducted by the first and second authors using a semi-structured interview guide (**Supplementary file D**). Interviews were conducted via the Teams^®^ digital meeting platform as a first option or a simple phone call if the participant so preferred or if internet connectivity was poor.

### Data management and analysis

Quantitative data were captured directly into an open-source database (REDcap^®^, Vanderbilt University). Data cleaning and analysis was carried out using Stata^®^ version 15 (College Station, Texas, USA). Summary statistics were compared across counties and multivariable modelling done using logistic regression. Systematicity in detection was assessed by correlating detection status with caller, pharmacy characteristics, and simulation process features.

Qualitative interviews were audio-recorded and transcribed verbatim in Microsoft Word^®^, translating into English where necessary. To ensure quality, all transcripts were cross-checked against the audio recordings. Transcripts were imported into NVivo 12 (SR International, Australia) and coded by the first and second authors. Analysis followed a thematic approach with initial categories based on the interview guide and emergent themes integrated in a second round of coding. A checklist on Consolidated criteria for Reporting Qualitative research (COREQ) was completed (**Supplementary file E**).

### Ethical considerations

The study was approved by the Kenya Medical Research Institute (KEMRI) Scientific and Ethics Review Unit (KEMRI/ SERU/CGMR-C/208/4080). Participants signed written informed consent forms administered digitally.

## Results

### Response rate and characteristics of participants

Of 195 target pharmacies, 108 (55%) completed a questionnaire. Participating pharmacies were generally similar to non-participating pharmacies, but had features indicative of higher operational capacity and interest in public health interventions, such as having a computerized stock management system, written job descriptions, and providing blood pressure measurement (data not shown). Ninety-four (87%) pharmacies were located in urban areas; 44 (41%) had a consultation room; 71 (67%) had a computerised stock management system; and 13 (12%) were part of a chain network **(Supplementary table I)**. The majority of questionnaire respondents were male, below 40 years, and had a pharmacy diploma.

One hundred and three (95%) pharmacies received a simulated client call. Thirteen (13%) calls were determined as probably detected. Detection was less likely if the call was made by one particular caller (perfect prediction), gender of person answering the call was male (adjusted odds ratio, aOR, 0.1, p=0.01); the pharmacy had a publicly available phone number (aOR 0.01, p=0.01), was part of a chain network (perfect prediction), opened 24 hours daily (perfect prediction), or had an on-site laboratory (perfect prediction). Main call outcomes (preventive therapies recommended, options for delivery of recommended products, and referral options) did not vary by detection status (data not shown), therefore all data were included in analysis.

Of 18 interview participants, 11 (61%) were male, 16 (88%) were below 40 years, 11 (61%) were pharmacy owner or in-charge, and 12 (67%) were diploma holders. Number of participants was balanced by county (6 each), but was skewed towards urban setting (13 v 5) and male gender (11 v 7) (**Supplementary Table II)**.

### Impact of the pandemic on service providers and pharmacy operations

Interview participants described fear and panic when they first heard about COVID-19, especially regarding the risk of exposure to infection:

> *“In the beginning, it sent everybody into a panic mode. Including my entire staff where everybody now thought this is a killer disease and being healthcare workers, they must be people who will be among the victims.”*
>
> - Male pharmaceutical technologist, Nairobi, 105

Some providers were in denial, feeling that the pandemic would never reach *“where we are”*, and others reported that some clients believed the pandemic was “*just an avenue maybe for our politicians ready to make some money or something”*.

The main concerns reported in the questionnaire were: exposure to infection at work (66% of respondents), “keeping my family safe” (22%), meeting financial needs (6%), and being caught up in the curfew (3%).

One hundred (93%) of 108 pharmacies reported pandemic-induced operational challenges (**Figure 1**). The main supply-side challenge was unavailability and high price of products and the main demand-side challenge was reduced client flow.

**Figure 1.**
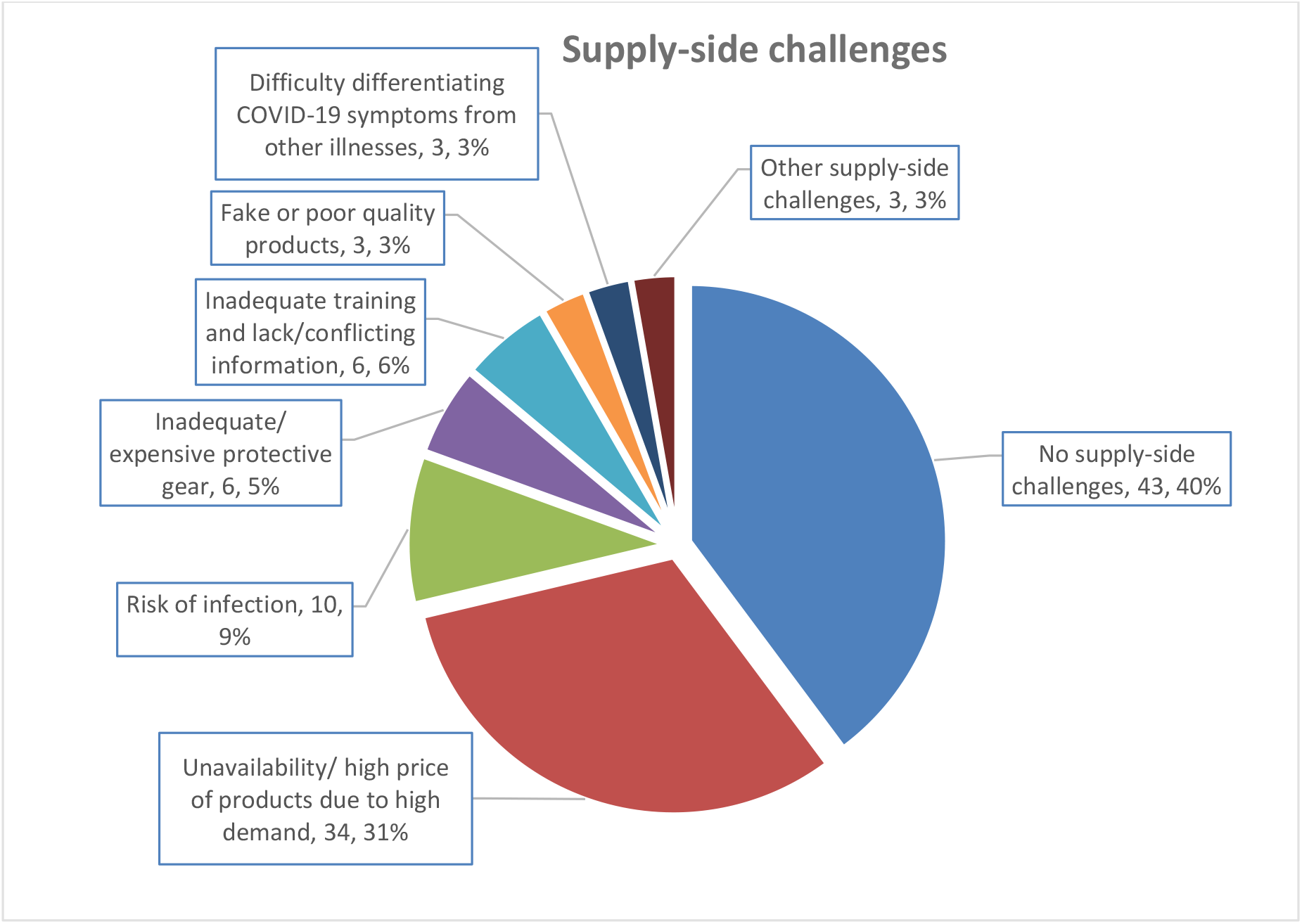
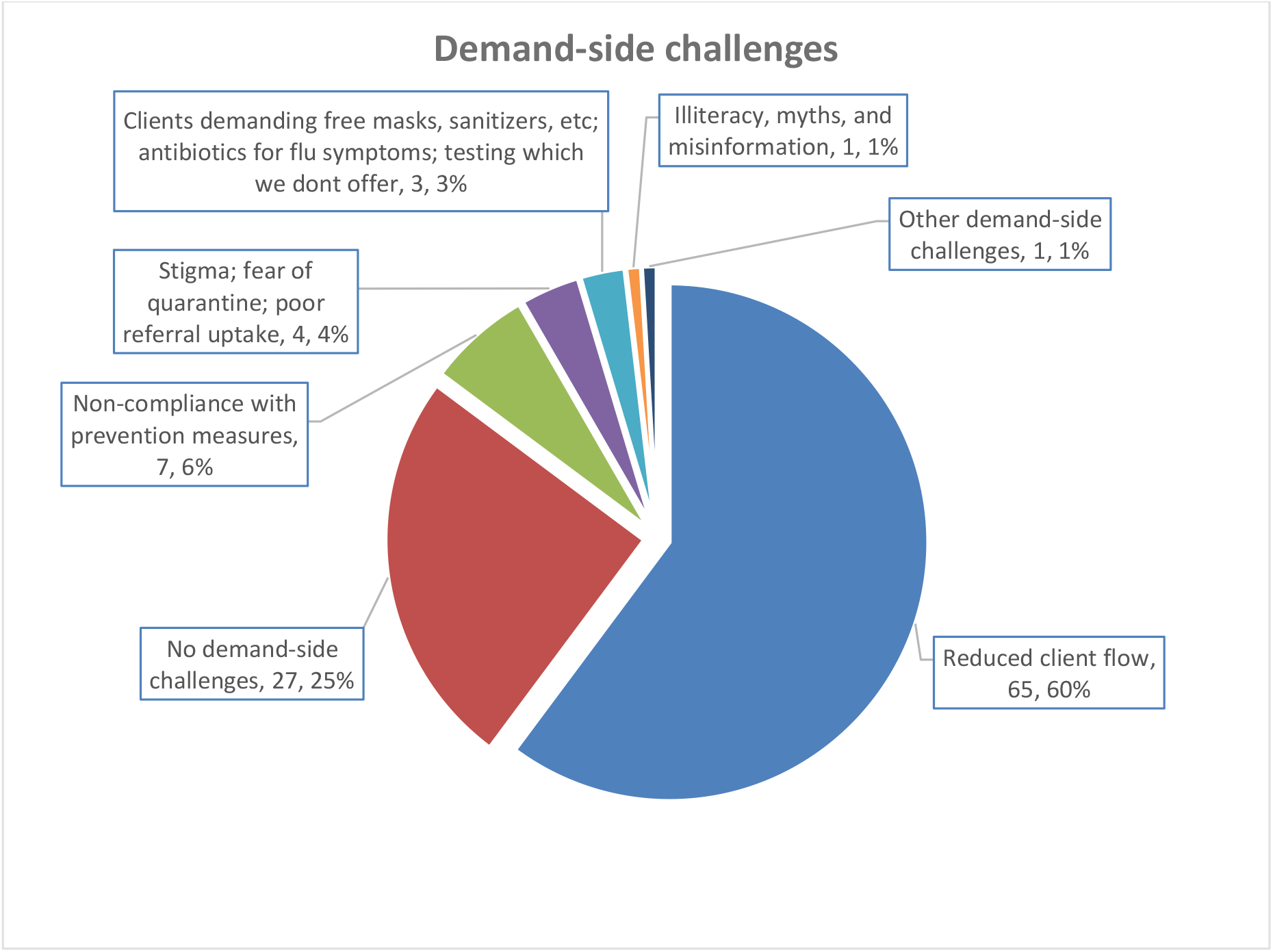
Challenges experienced by pharmacies during the COVID-19 pandemic in Kenya.

Interview data corroborated survey findings. Interviewees described temporal changes in client flow, with an initial surge of clients buying vitamins and other immune boosters, followed a few weeks later by a dip in demand to below pre-pandemic levels. The initial surge was accompanied by an increase in self-medication and stockpiling of long-term medications, such as for diabetes. While some of these requests were deemed potentially inappropriate, providers were not always able to effectively intervene:

> *“They just keep [these drugs] in case. …Like there was one [client] who came with a running nose, and then he was telling me he wanted Zithromax [a brand of the antibiotic azithromycin]. ‘Get Flu gone, get antihistamine, this one will help you’ [I told him]. So sometimes as a primary healthcare provider, you try to advise them, but now they have made up their mind, so you just go ahead and let them go [somewhere else]. Because whether you refuse, you do what, they’ve made up their mind.”*
>
> - Male pharmaceutical technologist, Kisumu, 303

Participants attributed the subsequent reduction in demand mainly to the dusk-to-dawn curfew, despite the fact that pharmacy providers were classified as essential workers and issued curfew passes:

> *“Of course, the curfew really beat us down, because now by the time [it’s] seven o’clock, people are going home… and it’s our prime time to serve clients as from 7 pm. So, we as well missed out on business.”*
>
> - Male pharmacy owner, Mombasa, 203

A number of interviewees believed that the decrease in service utilization was due to fear among clients, of infection and quarantine:

> *“Then people shied off from coming to the pharmacy especially the ones with coughs and flus because they were just thinking that I could tell them to go to hospital for the corona tests. So most clients decided to stay at home, not to come to the pharmacy for consultation, or they just sent somebody to buy them the drugs”*
>
> - Female pharmaceutical technologist, Kisumu, 304

One interviewee saw the stockpiling by clients in positive light, observing that it enabled him to offload slow-moving products:

> *“People resorted to scavenging where they might find these products and so that way [one was] able to liquidate what was lying in your shops”*
>
> - Male pharmaceutical technologist, Nairobi, 105

A reduction in hygiene-related illnesses was also hailed as a positive public health effect of the COVID-19 preventive measures:

> *“Due to proper maintenance of hygiene, the short-short illnesses have reduced. Washing of hands and stuff.””*
>
> - Male pharmacist, Nairobi, 103

### Adjustments to pharmacy operations

Pharmacies adjusted their operations to cope with these challenges and to accommodate the government-instituted prevention measures: 74% reduced opening hours (mean 3 hours [range 1-7]); 38% reduced staffing; and 30% changed their meeting routine (**Table 1**). Adjustments did not vary significantly across counties.

**Table 1.** Adjustments to pharmacy operations following the outbreak of COVID-19 in Kenya.

| Change | Nairobi<br>(N=38) |  | Mombasa<br>(N=36) |  | Kisumu<br>(N=34) |  | Total<br>(N=108) |  |
| --- | --- | --- | --- | --- | --- | --- | --- | --- |
|  | n | % | n | % | n | % | n | % |
| **Opening hours<sup>1</sup>** |  |  |  |  |  |  |  |  |
| No change | 7 | 18% | 8 | 22% | 10 | 29% | 25 | 23% |
| Reduced open hours per day | 31 | 82% | 25 | 69% | 24 | 71% | 80 | 74% |
| Reduced open days per week | 1 | 3% | 4 | 11% | 0 | 0% | 5 | 5% |
| Closed temporarily | 0 | 0% | 0 | 0% | 1 | 3% | 1 | 1% |
| **Staffing** |  |  |  |  |  |  |  |  |
| No change | 19 | 50% | 25 | 71% | 19 | 59% | 63 | 60% |
| Reduced number of staff | 18 | 47% | 9 | 26% | 13 | 41% | 40 | 38% |
| Increased number of staff | 0 | 0% | 1 | 3% | 0 | 0% | 1 | 1% |
| Reduced pay | 1 | 3% | 0 | 0% | 0 | 0% | 1 | 1% |
| **Stock volume levels** |  |  |  |  |  |  |  |  |
| No change | 12 | 32% | 13 | 36% | 8 | 24% | 33 | 31% |
| Reduced overall | 8 | 21% | 12 | 33% | 12 | 35% | 32 | 30% |
| Increased overall | 14 | 37% | 10 | 28% | 14 | 41% | 38 | 35% |
| Both reduced and increased for different items | 4 | 11% | 1 | 3% | 0 | 0% | 5 | 5% |
| **Meeting routine** |  |  |  |  |  |  |  |  |
| No change | 24 | 63% | 30 | 83% | 22 | 65% | 76 | 70% |
| Resorted to virtual/ remote/ online meetings<sup>2</sup> | 8 | 21% | 3 | 8% | 7 | 21% | 18 | 17% |
| Reduced frequency of meetings | 4 | 11% | 1 | 3% | 3 | 9% | 8 | 7% |
| Stopped meetings altogether | 2 | 5% | 1 | 3% | 0 | 0% | 3 | 3% |
| Reduced duration/ number of people | 0 | 0% | 1 | 3% | 1 | 3% | 2 | 2% |
| Changed timing | 0 | 0% | 0 | 0% | 1 | 3% | 1 | 1% |

<sup>1</sup> Three pharmacies in Mombasa and one in Nairobi reduced their open days per week by one, while one pharmacy in Mombasa reduced by three days. One pharmacy in Kisumu closed temporarily for 2 weeks
<sup>2</sup> Main virtual platforms used included Zoom, Google Meet, and Whatsapp

### COVID-19-related products and services provided by pharmacies

Table 2 presents the COVID-19-related products and services that were being provided or requested at pharmacies. Equipment and materials in highest demand were medical masks, N95/ K95 respirators, face shields, hand sanitizer, and thermometers; while the medications in highest demand were vitamin C, multivitamins, and antibiotics. Only 24 (22%) pharmacies reported keeping records related to COVID-19 services. Interview data suggested that many of these products and services were newly introduced following the outbreak, including products like vitamin C and surgical masks that one would have expected to find routinely in a pharmacy.

**Table 2.** COVID-19-related products and services provided by private retail pharmacies in Kenya. Data are number (%) of pharmacies providing or receiving requests (main row) and number of clients in past week (subsidiary row).

| Product/ service | Nairobi (N=38) |  | Mombasa (N=36) |  | Kisumu (N=34) |  | Total (N=108) |  |
| --- | --- | --- | --- | --- | --- | --- | --- | --- |
|  | N or median | % or IQR | N or median | % or IQR | N or median | % or IQR | N or median | % or IQR |
| **Equipment and materials** |  |  |  |  |  |  |  |  |
| Cloth masks | 1 | 3% | 5 | 14% | 6 | 18% | 12 | 11% |
| Number of clients in past week | 20 | 20-20 | 20 | 4-30 | 10 | 5-15 | 15 | 5-25 |
| Medical Masks | 37 | 97% | 33 | 92% | 30 | 88% | 100 | 93% |
| Number of clients in past week | 50 | 20-100 | 23 | 10-80 | 26 | 20-50 | 30 | 20-70 |
| N95/ K95 masks | 24 | 63%* | 18 | 50% | 11 | 32% | 53 | 49% |
|  | N or median | % or IQR | N or median | % or IQR | N or median | % or IQR | N or median | % or IQR |
| Number of clients in past week | 5 | 4-16 | 5 | 2-30 | 1 | 0-2 | 5 | 2-10 |
| Goggles | 6 | 16% | 2 | 6% | 3 | 9% | 11 | 10% |
| Number of clients in past week | 0 | 0-10 | 6 | 2-10 | 1 | 0-5 | 1 | 0-10 |
| Face shields | 19 | 50%* | 10 | 28% | 8 | 24% | 37 | 34% |
| Number of clients in past week | 1 | 0-6 | 1 | 0-3 | 1 | 0-8 | 1 | 0-5 |
| Gowns/ coveralls | 1 | 3% | 1 | 3% | 3 | 9% | 5 | 5% |
| Number of clients in past week | 0 | 0-0 | 10* | 0-10 | 0 | 0-1 | 0 | 0-1 |
| Hand sanitizer | 37 | 97% | 35 | 97% | 31 | 91% | 103 | 95% |
| Number of clients in past week | 10 | 3-30 | 5 | 2-13 | 5 | 2-13 | 5 | 3-15 |
| Thermometers | 27 | 71% | 23 | 64% | 20 | 59% | 70 | 65% |
| Number of clients in past week | 2 | 1-5 | 2 | 1-5 | 1 | 0-2 | 2 | 2-3 |
| Pulse oximeters | 10 | 26% | 11 | 31% | 3 | 9% | 24 | 22% |
| Number of clients in past week | 1 | 1-3 | 2 | 0-10 | 1 | 0-2 | 2 | 1-3 |
| **Medicines** |  |  |  |  |  |  |  |  |
| Vitamin C<sup>1</sup> | 35 | 92%* | 29 | 81% | 23 | 68% | 87 | 81% |
| Number of clients in past week | 10 | 10-30 | 15 | 10-30 | 5 | 3-11 | 10 | 4-20 |
| Multivitamins<sup>1</sup> | 29 | 76% | 30 | 83% | 24 | 71% | 83 | 77% |
| Number of clients in past week | 10 | 5-19 | 10 | 6-20 | 10 | 7-18 | 10 | 6-20 |
| Traditional/ alternative/ natural remedies<sup>1, 2</sup> | 1 | 3% | 2 | 6% | 0 | 0% | 3 | 3% |
| Number of clients in past week | 0 | 0-0 | 13 | 5-20 | 0 | 0-0 | 5 | 0-20 |
| Hydroxychloroquine/ chloroquine | 8 | 21% | 7 | 19% | 2 | 6% | 17 | 16% |
| Number of clients in past week | 3 | 2-7 | 1 | 0-5 | 3 | 0-5 | 2 | 0-5 |
| Antibiotics<sup>1, 3</sup> | 26 | 68% | 22 | 61% | 23 | 68% | 71 | 66% |
| Number of clients in past week | 3 | 2-10 | 15* | 10-39 | 15 | 5-29 | 10 | 3-20 |
| Steroids<sup>1</sup> | 16 | 42% | 16 | 44% | 16 | 47% | 48 | 44% |
| Number of clients in past week | 3 | 1-8 | 9 | 5-19 | 10 | 4-15 | 7 | 3-15 |
| Other medicines<sup>4</sup> | 3 | 8% | 4 | 11% | 2 | 6% | 9 | 8% |
| Number of clients in past week | 3 | 1-5 | 28 | 23-45 | 3 | 1-4 | 13 | 3-28 |
| **Information and counselling** |  |  |  |  |  |  |  |  |
| How to protect themselves from infection | 28 | 74% | 28 | 78% | 27 | 79% | 83 | 77% |
| Number of clients in past week | 5 | 2-13 | 5 | 3-15 | 10 | 3-20 | 5 | 3-15 |
| Symptoms to look out for if they suspect infection | 32 | 84% | 28 | 78% | 26 | 76% | 86 | 80% |
| Number of clients in past week | 3 | 2-10 | 5 | 4-10 | 9 | 2-11 | 5 | 2-11 |
| What to do if they develop symptoms/ suspect infection | 22 | 58% | 21 | 58% | 16 | 47% | 59 | 55% |
| Number of clients in past week | 3 | 2-7 | 3 | 2-5 | 5 | 3-22 | 4 | 2-10 |
| Mask use and related problems | 19 | 50% | 19 | 53% | 20 | 59% | 58 | 54% |
| Number of clients in past week | 5 | 2-10 | 5 | 2-35 | 15 | 4-20 | 7 | 2-20 |
| Counselling for anxiety related to getting infected/ becoming sick with COVID-19 | 14 | 37% | 16 | 44% | 13 | 38% | 43 | 40% |
| Number of clients in past week | 2 | 1-10 | 3 | 1-10 | 3 | 1-5 | 3 | 1-8 |
| Counselling for anxiety about job/ business loss | 5 | 13% | 7 | 19% | 5 | 15% | 17 | 16% |
|  | N or median | % or IQR | N or median | % or IQR | N or median | % or IQR | N or median | % or IQR |
| Number of clients in past week | 2 | 2-8 | 2 | 1-5 | 5 | 3-8 | 3 | 2-8 |
| **Modes of communication with clients** |  |  |  |  |  |  |  |  |
| Face-to-face at the pharmacy | 33 | 87% | 33 | 92% | 34 | 100% | 100 | 93% |
| Phone calls (including WhatsApp calls etc) | 21 | 55% | 25 | 69% | 20 | 59% | 66 | 61% |
| Texts (including WhatsApp messages, etc) | 13 | 34% | 15 | 42% | 12 | 35% | 40 | 37% |
| Facebook/ other web-based social media platforms | 3 | 8% | 2 | 6% | 2 | 6% | 7 | 6% |

*IQR: interquartile range*
*\* Indicates significant difference between counties ( $p < 0.05$ ), based on chi square test or analysis of variance for means*
*<sup>1</sup> A significant proportion of these sales may not have been related to COVID-19 prevention/ treatment*
*<sup>2</sup> Traditional/ alternative/ natural remedies included echinacea, chapa shoka, kaluma inhaler, menthol plus balm, and home remedies (lemon, papaya steaming)*
*<sup>3</sup> Antibiotics included azithromycin ( $n=55$ ), cephalosporin ( $n=7$ ), amoxicillin-containing ( $n=6$ ), other macrolide ( $n=2$ ); 3 pharmacies did not specify*
*<sup>4</sup> Other medicines included zinc+ vitamin D ( $n=4$ ), vitamin B complex ( $n=1$ ), salbutamol ( $n=1$ ), and scotts emulsion ( $n=1$ ); 2 pharmacies did not specify*

### Demand for COVID-19 testing and provider views towards pharmacy-based testing

Fifty-nine (55% [95% CI 45-65%]) of 108 pharmacies reported ever receiving a client asking for COVID-19 testing, with a median [interquartile range] number of requests in the last week of 2 [1-4]. At the time of the survey, none of the pharmacies was providing COVID-19 testing; and among the 59 pharmacies experiencing demand for testing, alternatives given to clients included referral to other testing sites (94%) and body temperature measurement (15%). Referral options included: public health facilities (92%), private health facilities (27%), and private labs (13%).

Interviews revealed that test requests were mostly by clients with symptoms, those who suspected they had been exposed, and those who needed certification for travel or employment purposes (eg, drivers and hotel staff).

When asked if they thought COVID-19 testing should be provided in pharmacies, 60% [95% CI 50-70%] said “yes”. Among the 64 that were supportive of pharmacy-based testing, main reasons for support were: pharmacies are easily accessible (36%), clients perceive pharmacies as more confidential (23%), clients shy away from health facilities (19%), and demand exists (9%).

In interviews, the confidentiality and convenience of the pharmacy setting was reiterated, with some interviewees suggesting home-based self-testing as an option:

> *“If it [testing] is available in pharmacies, somebody can just take the kit, you can just go home, you test yourself. If you find you are positive, you can just self-quarantine, without raising an alarm. So that when the quarantine period is over, you don’t get that kind of stigma, rejection.”*
>
> - Female pharmacy in charge, Mombasa, 201

Among the 42 that were not supportive of pharmacy-based testing, main reasons for opposing were: might be misused (29%), no private space in the pharmacy (24%), risk of infection (15%), protective gear is expensive or not available (12%), there is no demand for the service (10%), and staff are not adequately trained (10%). From the interviews, hesitancy to support pharmacy-based testing was mainly related to lack of capacity (*“I still need a thorough training…”*) and fear of infection.

### Handling of simulated clients seeking covid-19-related services

When a simulated client called a pharmacy to ask if they have medicines that the client can use to protect themselves from getting COVID-19, 76 (74%) of 103 pharmacies recommended one or more medications: 27% recommended at least one conventional therapy and 71% at least one alternative therapy or nutritional supplement (**Table 3**). The main recommendations given to a client suspecting COVID-19 infection were to: get tested, visit the nearest health facility, and self-quarantine at home.

**Table 3.** Handling of simulated clients seeking COVID-19-related services at private retail pharmacies in Kenya.

| Call outcome | Nairobi (N=38) |  | Mombasa (N=36) |  | Kisumu (N=34) |  | Total (N=103) |  |
| --- | --- | --- | --- | --- | --- | --- | --- | --- |
|  | n | % | n | % | n | % | n | % |
| **Provider response to “Do you have medicines that I can use to protect myself from getting COVID-19?”** |  |  |  |  |  |  |  |  |
| **Asked the client why he/she was asking for the medicines** | **14** | **38%\*** | **4** | **14%** | **2** | **6%** | **20** | **20%** |
| **Recommended conventional therapies** | **11** | **29%** | **6** | **19%** | **11** | **32%** | **28** | **27%** |
| Antibiotics | 11 | 29% | 6 | 19% | 10 | 29% | 27 | 26% |
| Azithromycin | 9 | 24% | 6 | 19% | 9 | 26% | 24 | 23% |
| Other antibiotics<sup>1</sup> | 2 | 5% | 1 | 3% | 4 | 12% | 7 | 7% |
| Painkillers | 1 | 3% | 0 | 0% | 3 | 9% | 4 | 4% |
| Antihistamines | 0 | 0% | 1 | 3% | 2 | 6% | 3 | 3% |
| **Recommended alternative therapies** | **29** | **76%** | **22** | **71%** | **22** | **65%** | **73** | **71%** |
| Vitamin C | 28 | 74% | 20 | 65% | 19 | 56% | 67 | 65% |
| Zinc | 25 | 66%* | 10 | 32% | 7 | 21% | 42 | 41% |
| Home remedies<sup>2</sup> | 6 | 16% | 4 | 13% | 6 | 18% | 16 | 16% |
| Vitamin B/ D | 4 | 11% | 4 | 13% | 2 | 6% | 10 | 10% |
| Multivitamins | 2 | 5% | 0 | 0% | 3 | 9% | 5 | 5% |
| Other remedies<sup>3</sup> | 1 | 3% | 0 | 0% | 1 | 3% | 2 | 2% |
| **Gave options on how to get the recommended medicines** | **37** | **97%** | **28** | **90%** | **34** | **100%** | **99** | **96%** |
| Pick from the pharmacy | 21 | 55% | 7 | 23%* | 18 | 53% | 46 | 45% |
| Home delivery | 19 | 50%* | 9 | 29% | 4 | 12% | 32 | 31% |
| Send someone to pick | 5 | 13% | 8 | 26% | 4 | 12% | 17 | 17% |
| **Provider response to “What symptoms should I look out for if I suspect COVID-19 infection?”** |  |  |  |  |  |  |  |  |
| **Mentioned any symptoms** | **34** | **89%** | **29** | **94%** | **30** | **88%** | **93** | **90%** |
| Fever | 23 | 61% | 20 | 65% | 21 | 62% | 64 | 62% |
| Cough | 13 | 34% | 15 | 48% | 16 | 47% | 44 | 43% |
| Flu-like symptoms | 15 | 39% | 15 | 48% | 12 | 35% | 42 | 41% |
| Breathing difficulty | 13 | 34% | 12 | 39% | 17 | 50% | 42 | 41% |
| Loss/ change of smell or taste | 16 | 42% | 13 | 42% | 11 | 32% | 40 | 39% |
| Sore throat | 15 | 39%* | 8 | 26% | 4 | 12% | 27 | 26% |
| Chest pain or congestion | 7 | 18% | 4 | 13% | 12 | 35%* | 23 | 22% |
| Headache | 6 | 16% | 4 | 13% | 8 | 24% | 18 | 17% |
| Fatigue, tiredness, malaise | 2 | 5% | 8 | 26%* | 4 | 12% | 14 | 14% |
| Reduced appetite | 2 | 5% | 2 | 6% | 7 | 21% | 11 | 11% |
| Body aches/ pains | 0 | 0% | 3 | 10% | 2 | 6% | 5 | 5% |
| Gut symptoms, eg, stomach pain, diarrhea, etc | 2 | 5% | 0 | 0% | 2 | 6% | 4 | 4% |
| Dizziness | 0 | 0% | 2 | 6% | 0 | 0% | 2 | 2% |
| Weight loss | 0 | 0% | 0 | 0% | 1 | 3% | 1 | 1% |
| **Provider response to “What should I do if I suspect I have COVID-19 infection?”** |  |  |  |  |  |  |  |  |
| **Immediate actions recommended** |  |  |  |  |  |  |  |  |
| Get tested for COVID-19 | 25 | 66% | 18 | 58% | 16 | 47% | 59 | 57% |
| Visit nearest health facility/ go see a doctor | 18 | 47% | 13 | 42% | 24 | 71%* | 55 | 53% |
| Self-quarantine/ isolate at home | 3 | 8% | 9 | 29%* | 4 | 12% | 16 | 16% |
| Take medications<sup>4</sup> | 3 | 8% | 2 | 6% | 3 | 9% | 8 | 8% |
| Check body temperature | 2 | 5% | 1 | 3% | 2 | 6% | 5 | 5% |
| Check oxygen levels | 0 | 0% | 1 | 3% | 1 | 3% | 2 | 2% |
| Check information online | 0 | 0% | 1 | 3% | 1 | 3% | 2 | 2% |
| Call 719<sup>5</sup> | 2 | 5% | 0 | 0% | 0 | 0% | 2 | 2% |
| **Supplementary actions recommended** |  |  |  |  |  |  |  |  |
| Wear a mask | 23 | 61% | 16 | 52% | 20 | 59% | 59 | 57% |
| Maintain physical distance | 21 | 55% | 15 | 48% | 18 | 53% | 54 | 52% |
| Observe hand hygiene | 7 | 18% | 5 | 16% | 7 | 21% | 19 | 18% |
| Eat fruits | 5 | 13% | 5 | 16% | 5 | 15% | 15 | 15% |
| Eat well<sup>6</sup> | 2 | 5% | 3 | 10% | 1 | 3% | 6 | 6% |
| Take lots of fluids | 1 | 3% | 1 | 3% | 3 | 9% | 5 | 5% |
| Live well<sup>7</sup> | 0 | 0% | 0 | 0% | 4 | 12% | 4 | 4% |
| Other advice<sup>8</sup> | 3 | 8% | 1 | 3% | 2 | 6% | 6 | 6% |
| **Caller assessment of the interaction** |  |  |  |  |  |  |  |  |
| How friendly (approachable) was the person who served you? |  |  |  |  |  |  |  |  |
| 1 - Very unfriendly | 1 | 3% | 0 | 0% | 0 | 0% | 1 | 1% |
| 2 - Unfriendly | 4 | 11% | 3 | 10% | 4 | 12% | 11 | 11% |
| 3 - Friendly | 20 | 53% | 22 | 71% | 22 | 65% | 64 | 62% |
| 4 - Very friendly | 13 | 34% | 6 | 19% | 8 | 24% | 27 | 26% |
| How enthusiastic was the provider about providing the service? |  |  |  |  |  |  |  |  |
| 1 - Very unenthusiastic | 1 | 3% | 0 | 0% | 0 | 0% | 1 | 1% |
| 2 - Unenthusiastic | 4 | 11% | 4 | 13% | 7 | 21% | 15 | 15% |
| 3 - Enthusiastic | 22 | 58% | 22 | 71% | 21 | 62% | 65 | 63% |
| 4 - Very enthusiastic | 11 | 29% | 5 | 16% | 6 | 18% | 22 | 21% |
| Did the provider say or do anything that you found particularly helpful or nice?<sup>9</sup> | 19 | 50% | 9 | 29% | 14 | 41% | 42 | 41% |
| Did the provider say or do anything that you found particularly NOT helpful or NOT nice?<sup>10</sup> | 4 | 11% | 2 | 6% | 7 | 21% | 13 | 13% |

\* Indicates significant difference between counties ( $p < 0.05$ ), based on chi square test
<sup>1</sup> Other antibiotics included Augmentin, Amoxil, ceftriaxone, etc
<sup>2</sup> Home remedies included ginger/ lemon/ hot water drink, hot soup/ water, garlic, steam, frequent hot baths
<sup>3</sup> Other alternative therapies included ginsomin and omega-3
<sup>4</sup> Medications for treatment included: azithromycin (3%), other antibiotics (4%), painkillers (2%), steroids (1%), home remedies (1%)
<sup>5</sup> 719 is the government hotline for COVID-19 reporting and information-seeking
<sup>6</sup> Eat well: eat healthy, eat vegetables, etc
<sup>7</sup> Live well: exercise, enough rest, fresh air, don't panic, avoid stress, etc
<sup>8</sup> Other advice: don't touch face, avoid handshakes, do not take any medicines, don't reuse mask, etc
<sup>9</sup> Things that were deemed particularly helpful or nice related mainly to: reassurance that infection risk is low if prevention measures are followed; high chance of recovery in case of infection; detailed information on what to do or where to seek further help; and follow-up calls in cases where the provider was busy at the time of the initial call.
<sup>10</sup> Things that were deemed not helpful or not nice related mainly to: calls being hanged up; inattentiveness or being in a hurry to finish the call; and dismissiveness (eg, "don't you watch news? The symptoms are public knowledge!" and "we don't have those medicines you are asking for, jaribu kwingine [try elsewhere])

### Infection prevention during service provision

The main infection prevention measures during service provision were hand hygiene, face masks, disinfection of services and physical distancing (**Table 4**). Eighty-five per cent of respondents rated the prevention measures instituted at their pharmacy as either adequate (72%) or very adequate (13%). Only one pharmacy reported that a staff member had ever tested positive for COVID-19, though no pharmacy reported regular testing of staff.

**Table 4.** COVID-19 infection prevention during service provision by private retail pharmacies in Kenya.

| Infection prevention measures | Nairobi (N=38) |  | Mombasa (N=36) |  | Kisumu (N=34) |  | Total (N=108) |  |
| --- | --- | --- | --- | --- | --- | --- | --- | --- |
|  | n | % | n | % | n | % | n | % |
| Among staff |  |  |  |  |  |  |  |  |
| Physical distancing<sup>1</sup> | 25 | 66% | 26 | 72% | 27 | 79% | 78 | 72% |
| Temperature monitoring | 13 | 34% | 14 | 39% | 13 | 38% | 40 | 37% |
| Hand hygiene | 35 | 92% | 33 | 92% | 32 | 94% | 100 | 93% |
| Disinfection of surfaces | 29 | 76% | 30 | 83% | 27 | 79% | 86 | 80% |
| Face masks<sup>2</sup> | 34 | 89% | 30 | 83% | 30 | 88% | 94 | 87% |
| Goggles | 0 | 0% | 1 | 3% | 1 | 3% | 2 | 2% |
| Ingestion of immune boosters | 1 | 3% | 0 | 0% | 0 | 0% | 1 | 1% |
| Among clients |  |  |  |  |  |  |  |  |
| Physical distancing<sup>1</sup> | 33 | 87% | 31 | 86% | 30 | 88% | 94 | 87% |
| Temperature monitoring | 13 | 34% | 9 | 25% | 11 | 32% | 33 | 31% |
| Hand hygiene<sup>3</sup> | 36 | 95% | 33 | 92% | 30 | 88% | 99 | 92% |
| Face masks | 27 | 71% | 32 | 89% | 27 | 79% | 86 | 80% |
| Non-touch payment methods<sup>4</sup> | 14 | 37% | 16 | 44% | 10 | 29% | 40 | 37% |
| Information materials and messages<sup>5</sup> | 12 | 32% | 13 | 36% | 11 | 32% | 36 | 33% |

<sup>1</sup> Physical distancing was achieved through various means including: reducing number of staff and working in shifts, ensuring a distance of 1.5 -3 meters between staff and clients through the use of posters and floor/seat markings, serving one client at a time or other crowd control measures, ribbons/ ropes/ glass/ metal-grill/ wooden barriers between client and provider, and home delivery of products. One pharmacy noted that “patients with cough and related symptoms are always first referred to the health center for covid19 testing before being served further”.
<sup>2</sup> Majority of pharmacies (95%) were using surgical/medical masks; only 6% reported use of cloth masks.
<sup>3</sup> Hand hygiene measures among clients included: handwashing, hand sanitizer, and “no hand-shakes” policy
<sup>4</sup> Non-touch payment methods included: mobile money, and credit/ debit cards; one pharmacy reported disinfection of cash.
<sup>5</sup> Messages were delivered through: posters (21%), leaflets (5%), television screens (2%), and audio announcements (1%)

### Information sources and training

Sources of general COVID-19 information included: continuing professional development sessions (37% of respondents), ministry of health (MoH) communications (23%), social media and internet (19%), television and radio (12%), Kenya Pharmaceutical Association (KPA)/ Pharmaceutical Society of Kenya (PSK) communications (5%), scientific journals (2%) and colleagues (2%). For specific questions about COVID-19, providers consulted the pharmacy-in-charge (48%), nearby doctor (37%), MoH officials (32%), the internet (5%), and medical representatives (2%).

Interviewees felt that the official announcement of the first case played was a major source of awareness, though some reported having learnt about the outbreak through radio, television, and social media prior to the first reported case. However, they decried the rapidly changing and sometimes contradictory information, for example *“on whether to use or not to use masks”*.

Fifty-two (48%) pharmacies had at least one staff member trained on COVID-19, with a median [range] of 1 [1-8] providers trained. This did not vary by county or by membership in professional associations. Modalities of training included: on-site in the pharmacy (67% of respondents), virtual (54%), and off-site (13%). Virtual trainings were more prevalent in Kisumu compared to Nairobi (76% vs 35%, p=0.04). Training sessions were facilitated by MoH (73%), KPA (51%), PSK (19%), AMREF (9%), and Kenyatta National Hospital (5%). While some interview participants felt they had received “a lot’’ of training, others felt the trainings were not very adequate, particularly with regard to inclusion of updated information in follow-up sessions and the fact that most trainings included only the pharmacy in-charge and not the entire staff.

### External support and linkages

Interviewees expressed appreciation for guidance from the ministry of health, including the pharmacy and poisons board, who “*did some inspection and brought some banners [information materials] on how I can protect myself and the general public.”* The government-led community awareness-creation activities were also hailed as an important complement to pharmacy efforts in educating the public.

However, examples of direct linkages and material support were rare. One interviewee in Nairobi reported that community health workers from a nearby health centre regularly visited the pharmacy to collect information on number of suspect cases seen. Another interviewee in Kisumu described an instance where an ambulance was sent promptly on reporting a patient in critical condition.

More commonly, a feeling of isolation and neglect prevailed, with pharmacies reporting that they were not receiving much support from the government or other external parties with regard to COVID-19, unlike what happens with other public health programs:

> *“So far being a private facility, we are not receiving any financial support from the organizations in COVID related issues. We don’t receive that apart from PS Kenya [NGO supporting TB and HIV programs].”*
>
> - Male pharmaceutical technologist, Kisumu, 301

### Confidence, motivation, and coping mechanisms

Majority of questionnaire respondents reported feeling confident (68%) or very confident (19%) about providing COVID-19-related services. Over half felt prepared (51%) or very prepared (7%) about handling a client who presents to the pharmacy with COVID-19 symptoms.

Motivation for supporting delivery of COVID-19 services included: “to help people protect themselves or prevent transmission (75%), “It’s part of my job/ I am required to do it” (14%), encouragement and support from professional associations (7%), “to boost the profile of the pharmacy” (2%), and “to make money” (1%). When asked how much they felt their personal role in the COVID-19 response was valued by the community, the majority stated highly valued (68%) or very highly valued (11%)

In addition to the operational adjustments described above, interviewees described other coping mechanisms such putting up a shade outside to prevent crowding inside the pharmacy and improvisation of hand sanitizers when not available from suppliers. Some interviewees also reported going the extra mile sometimes, eg, by providing free masks to clients.

### Provider suggestions and recommendations

Seventy-six (70%) questionnaire respondents highlighted inputs that they felt pharmacies can uniquely contribute to the national response, including: provision of information and counselling to clients (64%), distribution of preventive materials (21%), triage and referral of suspect cases (17%), surveillance of infection rates and community adherence to preventive measures (14%), provision of preventive and curative therapies (12%), and maintenance of long-term therapy (3%). Pharmacies’ location within communities was cited as a strength when it comes to surveillance of client behaviour and community awareness creation.

To facilitate these inputs, pharmacies suggested additional support that they would require from the government and other external parties, including: training (34% of respondents), free or subsidized preventive materials such as masks, sanitizers, and thermo guns for own use and distribution to clients (34%), supply management such as price and quality controls (15%), clear guidelines and referral systems (9%), community awareness creation (7%), information materials (6%), financial support such as loan repayment holidays and medical insurance (6%), and testing services (5%).

These suggestions were echoed in interviews. Post-marketing surveillance to ensure quality of products in the market and a streamlined process to capture and transmit information back to the government were pinpointed as key gaps in the pandemic response. Overall, there were earnest calls for more deliberate engagement, oversight, and support supervision by the government and other implementing partners:

> *“If the government could put us, the pharmacists and pharm techs in the you know, treat us like people who are fighting this virus and facilitate this thing it would be a major shift, it would be a major shift.”*
>
> - Male pharmacy in charge, Mombasa, 205

## Discussion

In this study among private retail pharmacies (community pharmacies) in Kenya, we describe the impact of the COVID-19 pandemic on operations, COVID-19-related products and services being provided, infection prevention during service prevention, and provider suggestions and recommendations on how to improve pandemic preparedness and response.

The initial weeks after the pandemic reached Kenya were characterized by fear and panic among providers and a surge in client flow. Broadly speaking, the experiences documented in our study can be captured with the apt words of Austin et al in their paper describing the experiences of community pharmacists in Ontario, Canada: “the scale and magnitude of the COVID-19 pandemic defied imagination or pre-planning, yet health care professionals like pharmacists were required to continue to provide services and care to patients – and in many cases, expand their repertoire of clinical skills to assume new and even more challenging responsibilities” [15].

The initial surge in demand waned rapidly, with client flow dipping below pre-pandemic levels within a few weeks and remaining depressed until the time of data collection, seven months later. This was surprising as we had hypothesized, before the study, that pharmacies would experience an overall increase in demand as patients redirected their care-seeking from health facilities for fear of exposure to infection and quarantine. Participants attributed the decrease in demand mainly to the movement restrictions instituted by government. However, they also suggested – ironically – that clients, especially those with flu-like symptoms, may have avoided pharmacies for fear of being referred to public facilities and subsequently being quarantined.

Our findings clearly show that community pharmacies in Kenya were undertaking potentially impactful activities to support the response to the COVID-19 pandemic. Despite a feeling of disconnection from the government-led national response, providers expressed great readiness and motivation for provision of COVID-19-related services, similar to providers in Qatar [30], the USA [31], and other settings around the world [32]. The services pharmacies in our study were providing are generally similar to those suggested by experts in the field [22, 33-35], as well as those being provided in other low-to-middle income countries [36-39] and in high-income settings [30, 40, 41].

However, management of drug shortages and prevention of stockpiling (hoarding) by clients [22, 30, 33] were reported only as challenges and not as roles that Kenyan pharmacies were playing or could play. Additionally, while 60% of questionnaire respondents said they were receiving queries and providing information and counseling over the phone, other remote services such as online prescribing and home delivery of medications [22, 36] were not being widely used or mentioned as potential additional roles. Participants in our study, being frontline workers, may have deemed the carrying out of these innovations as being beyond the sphere of their influence. Similarly, COVID-19 vaccine administration was not mentioned as a potential role, understandably perhaps because a vaccine was not available at the time. But pharmacies in Kenya are actively involved in provision of other vaccines [42, 43], hence it is reasonable to hypothesize that COVID-19 vaccines could be delivered through private pharmacies in the future. Interviews with policymakers and captains of industry may be required to probe these topics further.

The finding that no pharmacy was providing COVID-19 testing was unsurprising since no approved point-of-care (POC) tests were available at the time of the study. The more interesting finding is that there was significant demand and support for pharmacy-based testing, similar to what studies in the Middle East [44] and the USA [31] have found. In this context, and given the fact that at least three rapid POC tests have since been approved in Kenya [45], there is an urgent need to start developing policies to guide implementation. This will obviate the emergence of an uncoordinated and potentially dangerous service, similar to the HIV testing one that we observed in 2016 before policy guidelines were issued [46, 47].

For treatment of COVID-19 symptoms, most pharmacies recommended alternative therapies and nutritional supplements such as vitamin C and zinc, and only about a third suggested conventional therapies with no proven efficacy such as antibiotics [13, 14]. This finding is similar to that of a small study in Nairobi in May 2020 which found that despite increased requests for antimalarials and antibiotics, pharmacists recommended alternative therapies [48]. A similar finding was obtained in a multinational survey of Asian pharmacists [35]. While alternative therapies and nutritional supplements are themselves not evidence-based (as no rigorous trials have proven their efficacy against SARS-CoV-2), they have a better safety profile and their useful effects are more biologically plausible since they have been used for ages against the flu virus, itself a coronavirus. These findings are therefore encouraging. However, among Egyptian pharmacies, the antibiotic azithromycin was given to about 40% of presumptive COVID-19 patients with mild to moderate symptoms, albeit based on physician recommendation [49]. Further research in different jurisdictions may be needed to describe and contrast the use of antibiotics and other unproven COVID-19 therapies, in order to inform the promotion of rational drug use and the prevention of antimicrobial resistance.

Our study identified other important gaps in community pharmacies’ preparedness and response to the pandemic. Notably, pharmacies did not seem to have any well-defined operational linkages to other units of the healthcare system, including for example, the emergency operations center at the ministry of health. Besides, while only about half of pharmacies had at least one staff member trained on COVID-19, some of those trained felt they had received a lot of training. This suggests a “network effect” that our study was unable to decipher, where a certain group of providers seem to be well-positioned to receive many trainings, while others receive none.

Specific programs will be required to address these gaps and enhance pandemic preparedness. Templates for public-private cooperation in pandemic responses do exist from other settings. Ung *et al*, for example, report how in one administrative region on the south-east coast of China, pre-existing public-private partnerships between the local government and community pharmacies ensured rapid provision and prevented hoarding of face masks through a government-led “Guaranteed Mask Supply for Macao Residents Scheme” [50]. A qualitative study among pharmacists in Canada, identified key features that predicted community pharmacists’ resilience during the early period of the pandemic, including: use of and comfort with technology; early adoption of corporate and professional guidance; emphasis on task-focus rather than multi-tasking; and provision of personal protective equipment [15]. Roll-out of pharmacy-based COVID-19 programs should be accompanied with implementation studies to better uncover aspects of capacity, preparedness, and resilience.

Our study had a number of strengths. First, we took advantage of information collected in a recent study to speed up recruitment and data collection. Second, we used multiple quantitative and qualitative methods to enhance the comprehensiveness and reliability of the findings. Of note, we used simulated clients to objectively assess provider practices, an approach that is gaining acceptance as a gold standard for the measurement of clinical practice quality around the world [51]. One weakness of our study was the low response rate (55%), considering that we targeted pharmacies that had participated in a similar study less than a year previously. Compared to non-participating pharmacies, participating pharmacies had features suggestive of higher operational capacity and interest in public health interventions. While this limits generalizability of our findings, it can also be looked at in positive light. We propose that pharmacies with these characteristics could be identified for inclusion in a “practice and learning network” that could provide a platform for testing implementation strategies for various public health interventions [52-54]. Scale up of such interventions would then target similar pharmacies and include strategies for enhancing operational capacity and interest in public health among the other pharmacies.

In conclusion, we found that private retail pharmacies in Kenya were actively contributing to the COVID-19 response despite an apparent disconnection from the national response program. More deliberate engagement, support and linkages are required. Notably, there is an urgent need to develop policy guidelines for pharmacy-based COVID-19 testing, a service that is clearly needed and which could greatly increase test coverage. Roll-out of this and other pharmacy-based COVID-19 programs should be accompanied with implementation research in order to inform current and future pandemic responses.

## Supporting information

Supplementary Table I. Survey participants

Supplementary Table II. Inteview participants-2

## Data Availability

All data produced in the present study are available upon reasonable request to the authors

## Acknowledgments

We acknowledge the contributions of all participating pharmacy service providers. We thank James Wafula for supporting database development and data management; and Langat Justus, Lucy Omumbo, Milkah Auka, and Rodgers Ayoma for supporting the simulated client survey.

## Author Contributions

Conceived the study: PM EB. Developed protocol: PM DM SM EB. Performed the experiments: PM AM; Analysed the data: PM AM JN. Wrote the manuscript: PM AM DM JN SM EB. Approved the final manuscript: PM AM DM JN SM EB.

The authors declare not conflicts of interest.

## Patient and Public Involvement

Findings will be shared with the pharmacy sector at the 2021 Pharmaceutical Society of Kenya Annual Conference and a policy brief will be prepared targeting stakeholders in the ministry of health, pharmacy sector and other implementing partners.

