## Supplementary Table I. Survey participants for "Response to the coronavirus disease 2019 (COVID-19) pandemic at private retail pharmacies in Kenya: a mixed methods study"

**Supplementary Table I: Characteristics of participating pharmacies and respondents**

*A. Characteristics of participating pharmacies*

| Characteristic | Nairobi (n=38) | | Mombasa (n=36) | | Kisumu (n=34) | | Total (n=108) ^ | |
|---|---|---|---|---|---|---|---|---|
| | n | % | n | % | n | % | n | % |
| **Setting** | | | | | | | | |
| Urban commercial centre | 23 | 61% | 29 | 81% | 12 | 35% | 64 | 59% |
| Urban residential area | 13 | 34% | 4 | 11% | 6 | 18% | 23 | 21% |
| Urban informal settlement | 2 | 5% | 3 | 8% | 2 | 6% | 7 | 6% |
| Rural town/ shopping centre | 0 | 0% | 0 | 0% | 14 | 41%* | 14 | 13% |
| **Years of operation** | | | | | | | | |
| Number of years in operation (median [range]) | 5 | 1-19 | 4 | 1-42 | 5 | 1-11 | 5 | 1-42 |
| **Physical facilities** | | | | | | | | |
| Total number of rooms (Median [range]) | 2 | 1-10 | 2 | 1-6 | 2 | 1-8 | 1 | 1-10 |
| Consultation room | 14 | 37% | 10 | 28% | 20 | 59%* | 44 | 41% |
| On-site lab | 5 | 13% | 1 | 3% | 6 | 18% | 12 | 11% |
| **Information systems** | | | | | | | | |
| Computerized stock system | 29 | 81%* | 19 | 53% | 23 | 68% | 71 | 67% |
| Individual medical records[1] | 9 | 25% | 2 | 6% | 7 | 21% | 18 | 17% |
| **Chain network membership[2]** | | | | | | | | |
| Part of a chain network | 9 | 24%* | 3 | 8% | 1 | 3% | 13 | 12% |
| Number of branches nationwide (Median [range]) | 1 | 1-41 | 1 | 1-41 | 1 | 1-9 | 1 | 1-41 |
| **Branding[3]** | | | | | | | | |
| None | 33 | 87% | 29 | 81% | 32 | 94% | 94 | 87% |
| Green Cross | 2 | 5% | 2 | 6% | 0 | 0% | 4 | 4% |
| Pharmnet (blue cross) | 0 | 0% | 3 | 8% | 1 | 3% | 4 | 4% |
| Corporate | 3 | 8% | 2 | 6% | 1 | 3% | 6 | 6% |
| **Distance to the nearest clinic** | | | | | | | | |
| Within the pharmacy | 0 | 0% | 0 | 0% | 2 | 6% | 2 | 2% |
| < 100 meters | 17 | 45% | 13 | 36% | 9 | 26% | 39 | 36% |
| < 1km | 19 | 50% | 16 | 44% | 21 | 62% | 56 | 52% |
| < 5km | 2 | 5% | 7 | 19% | 2 | 6% | 11 | 10% |
| **Distance to the nearest HIV testing site** | | | | | | | | |
| < 100 meters | 14 | 37% | 12 | 33% | 11 | 32% | 37 | 34% |
| < 1km | 18 | 47% | 16 | 44% | 21 | 62% | 55 | 51% |
| < 5km | 4 | 11% | 7 | 19% | 2 | 6% | 13 | 12% |
| >5km | 1 | 3% | 0 | 0% | 0 | 0% | 1 | 1% |
| Not sure | 1 | 3% | 1 | 3% | 0 | 0% | 2 | 2% |
| **Service providers** | | | | | | | | |
| Total number of service providers (Median [range]) | 3* | 1-16 | 2 | 1-9 | 2 | 1-4 | 2 | 1-16 |
| At least one pharmacy degree holder | 8 | 21% | 7 | 19% | 3 | 9% | 18 | 17% |

| Characteristic | Nairobi (n=38) | | Mombasa (n=36) | | Kisumu (n=34) | | Total (n=108) ^ | |
|---|---|---|---|---|---|---|---|---|
| | n | % | n | % | n | % | n | % |
| At least one pharmacy diploma holder | 36 | 95% | 35 | 97% | 34 | 100% | 105 | 97% |
| All staff belong in a professional association | 24 | 63% | 19 | 53% | 18 | 53% | 61 | 56% |
| **Management** | | | | | | | | |
| Written job descriptions | 21 | 55% | 12 | 33% | 14 | 41% | 47 | 44% |
| Regular staff meetings[4] | 21 | 55% | 16 | 44% | 19 | 56% | 56 | 52% |
| **Opening hours** | | | | | | | | |
| 24 hours daily, 7 days a week | 4 | 11% | 1 | 3% | 1 | 3% | 6 | 6% |
| At least seven days a week | 23 | 61% | 28 | 78% | 25 | 74% | 76 | 70% |
| At least 6 days a week | 38 | 100% | 34 | 94% | 34 | 100% | 106 | 98% |
| **Work load** | | | | | | | | |
| Walk-in clients per day (Median [range]) | 55 | 15-300 | 50 | 20-200 | 40 | 20-200 | 50 | 15-300 |
| Online clients per day (Median [range]) | 1 | 0-20 | 0 | 0-20 | 0 | 0-10 | 0 | 0-20 |
| **Health promotion services** | | | | | | | | |
| Over-the-counter STI treatment | 29 | 76% | 29 | 81% | 30 | 88% | 88 | 81% |
| Weight management | 12 | 32% | 6 | 17% | 10 | 29% | 28 | 26% |
| Smoking cessation | 4 | 11% | 1 | 3% | 1 | 3% | 6 | 6% |
| Vaccination | 0 | 0% | 1 | 3% | 3 | 9% | 4 | 4% |
| **Screening services** | | | | | | | | |
| Blood pressure measurement | 26 | 68% | 26 | 72% | 25 | 74% | 77 | 71% |
| Blood sugar testing | 19 | 50% | 25 | 69% | 20 | 59% | 64 | 59% |
| HIV self-testing | 21 | 55% | 22 | 61% | 20 | 59% | 63 | 58% |
| Malaria testing | 9 | 24% | 19 | 53% | 27 | 79%* | 55 | 51% |
| In-pharmacy pregnancy testing | 9 | 24% | 9 | 25% | 18 | 53%* | 36 | 33% |
| Other screening services[5] | 1 | 3% | 0 | 0% | 5 | 15%* | 6 | 6% |

*^ Of 195 target pharmacies that had participated in the previous PHP study, 108 (55%) participated in the current survey. Of 87 that did not participate: 55 (63%) failed to complete the questionnaire after three reminders, 15 (17%) reported that the initial respondent had left and no one else was willing to participate, 8 (9%) were unreachable, 3 (3%) reported that the management did not approve participation, 2 (1%) were undergoing transition, 1 (1%) was closed, and 3 (3%) had other reasons (no access to smartphone or computer, preferred face-to-face interview, sold the pharmacy).*

** Indicates significant difference between counties (p<0.05), based on chi square test or analysis of variance for means*

*[1] An individual medical record is a document detailing the medications an individual patient is on, indications and drug allergies; may also include other clinical information.*

*[2] Chain network was defined as 6 or more branches nationwide, derived from chain size distribution pattern; 79 (73%) pharmacies were single branch, 16 (15%) were part of a 2-5 branch network, and 13 (12%) were part of a chain network*

*[3] Green Cross is a professional network of pharmacies owned by pharmacists; Pharmnet is for pharmaceutical technologists*

*[4] Excludes 18 pharmacies that responded "not applicable" since they have only one staff member*

*[5] Other screening services included; typhoid testing (n=4) and tuberculosis screening (n=2)*

*STI: Sexually transmitted infection*

*B. Characteristics of respondents*

| Characteristic | Nairobi (N=38) | | Mombasa (N=36) | | Kisumu (N=34) | | Total (N=108) | |
|---|---|---|---|---|---|---|---|---|
| | n | % | n | % | n | % | n | % |
| **Gender** | | | | | | | | |
| Female | 12 | 32% | 13 | 36% | 16 | 47% | 41 | 38% |
| Male | 26 | 68% | 23 | 64% | 18 | 53% | 67 | 62% |
| **Age** | | | | | | | | |
| 20 – 29 years | 12 | 32% | 12 | 33% | 14 | 41% | 38 | 35% |
| 30 – 39 years | 19 | 50% | 18 | 50% | 16 | 47% | 53 | 49% |
| 40 – 44 years | 5 | 13% | 4 | 11% | 2 | 6% | 11 | 10% |
| 45 years and over | 2 | 5% | 2 | 6% | 2 | 6% | 6 | 6% |
| **Designation (role)** | | | | | | | | |
| Owner | 9 | 24% | 12 | 33% | 12 | 35% | 33 | 31% |
| In-charge/ Superintendent | 11 | 29% | 13 | 36% | 8 | 24% | 32 | 30% |
| Staff[1] | 18 | 47% | 11 | 31% | 14 | 41% | 43 | 40% |
| **Highest level of education[2]** | | | | | | | | |
| Non-health qualifications | 0 | 0% | 2 | 6% | 1 | 3% | 3 | 3% |
| Health-related certificate/ diploma/ bachelor's degree | 2 | 5% | 0 | 0% | 2 | 6% | 4 | 4% |
| Pharmacy certificate | 2 | 5% | 0 | 0% | 1 | 3% | 3 | 3% |
| Pharmacy Diploma | 29 | 76% | 31 | 86% | 28 | 82% | 88 | 81% |
| Pharmacy degree | 3 | 8% | 3 | 8% | 0 | 0% | 6 | 6% |
| Health-related master's degree | 2 | 5% | 0 | 0% | 2 | 6% | 4 | 4% |
| **Experience (Median [Range])** | | | | | | | | |
| Total number of years worked | 7 | 1-25 | 5 | 2-41 | 6 | 2-20 | 6 | 1-41 |
| Number of years worked in the current pharmacy | 3 | 1-14 | 3 | 1-30 | 3 | 1-17 | 3 | 1-30 |

*[1] Staff designations included: pharmaceutical technologist (n=35), pharmacist (n=5), pharmacy assistant (n=2), and nurse aid (n=1)*
*[2] Non-health qualifications included: business management (n=1), supplies management (n=1), and college (n=1); health-related certificate/ diploma/ bachelor's degree included: medicine (n=1), nursing (n=2), and environmental health (n=1)h; health-related master's degree included: pharmacy (n=1), and other (n=3)*
