## Supplementary Table II. Inteview participants-2 for "Response to the coronavirus disease 2019 (COVID-19) pandemic at private retail pharmacies in Kenya: a mixed methods study"

**Supplementary Table II. Characteristics of in-depth interview participants.** The interviewee # is used to reference illustrative quotes in the results section. Rows are sorted by interviewee #.

| Interviewee # | County | Practice setting | Sex | Age category (years) | Highest level of education | Years in practice | Position |
|---|---|---|---|---|---|---|---|
| 101 | Nairobi | Urban residential area | Female | 20 - 24 | Pharmacy Diploma | 4 | Staff |
| 102 | Nairobi | Urban commercial center | Male | 30 - 34 | Pharmacy Diploma | 10 | Superintendent |
| 103 | Nairobi | Urban commercial center | Male | 25 - 29 | Pharmacy degree | 2 | Staff |
| 104 | Nairobi | Urban commercial center | Female | 30 - 34 | pharmacy assistant certificate | 8 | Staff |
| 105 | Nairobi | Urban commercial center | Male | 25 - 29 | Pharmacy Diploma | 6 | Superintendent |
| 106 | Nairobi | Urban residential area | Male | 35 - 39 | Pharmacy Diploma | 10 | Superintendent |
| 201 | Mombasa | Urban commercial center | Female | 25 - 29 | Pharmacy Diploma | 5 | In-charge |
| 202 | Mombasa | Urban commercial center | Male | 40 - 44 | Pharmacy Diploma | 10 | Owner or director |
| 203 | Mombasa | Urban informal settlement | Male | 35 - 39 | BSc supply chain | 5 | Owner or director |
| 204 | Mombasa | Urban commercial center | Male | 25 - 29 | Pharmacy Diploma | 4 | Staff |
| 205 | Mombasa | Urban commercial center | Male | 25 - 29 | Pharmacy Diploma | 4 | Superintendent |
| 206 | Mombasa | Urban commercial center | Female | 20 - 24 | Pharmacy Diploma | 2 | Staff |
| 301 | Kisumu | Urban commercial center | Male | 25 - 29 | Pharmacy Diploma | 2 | Staff |
| 302 | Kisumu | Rural town/ shopping center | Female | 40 - 44 | Nursing certificate | 17 | Staff |
| 303 | Kisumu | Urban commercial center | Male | 30 - 34 | Pharmacy Diploma | 7 | Superintendent |
| 304 | Kisumu | Rural town/ shopping center | Female | 35 - 39 | Pharmacy Diploma | 7 | Superintendent |
| 305 | Kisumu | Rural town/ shopping center | Female | 30 - 34 | Health-related masters | 9 | Owner or director |
| 306 | Kisumu | Rural town/ shopping center | Male | 35 - 39 | Health-related masters | 10 | Owner or director |
