## Supplementary files A-E for "Response to the coronavirus disease 2019 (COVID-19) pandemic at private retail pharmacies in Kenya: a mixed methods study"

**<u>Supplementary file A – PHARMACY QUESTIONNAIRE</u>**

**Note to interviewer:** This questionnaire is targeted at the pharmacy service provider (PSP) previously included in the PHP study. In case they are not available, target the in-charge or most experienced or most highly-educated PSP, in that order.

**BASIC INFORMATION**
Interviewer identifier; Pharmacy identifier; Date; Time

**INTRODUCTION**
*Thank you for finding time for this interview. I would like to ask you some questions regarding general operations in your pharmacy and your experience providing COVID19-related services…*

**<u>SECTION I: RESPONDENT CHARACTERISTICS</u>**

1. What is your gender? Male/Female/other, specify
2. What is your age category in years?
   - 18 – 24
   - 25 – 29
   - 30 – 34
   - 35 – 39
   - 40 – 44
   - 45 – 49
   - 50 – 54
   - 55 – 59
   - >59
3. What position do you hold in the pharmacy? [pharmacy owner or director/ Superintendent/ Pharmacy in-charge/ Pharm Tech/ Pharmacist/ other, specify]
4. What is your highest level of education? Pharmacy degree/ Pharmacy Diploma / Other
5. How many years have you worked in the retail/ community pharmacy sector? [numeric; if less than a year, enter 0.5]
   a. How many years have you worked in the current pharmacy?

**<u>SECTION II: OVERALL EFFECT OF THE COVID19 EPIDEMIC ON PHARMACY OPERATIONS</u>**

6. Have you had to make any changes to your operations since the outbreak of COVID19 epidemic in Kenya (mid-March 2020)? Yes/no
   a. Have you had to change your hours of operation? [can select more than one]
      - No
      - Reduced open hours per day, specify number of hours reduced
      - Reduced open days per week, specify number of days reduced
      - Closed for some time then re-opened, specify closure period in weeks
      - Closed indefinitely, specify date of closure
      - Other, specify
   b. Have you had to change number of staff? [select one]
      - No
      - Reduced
      - Increased
      - Other, specify
   c. Have you had to change stocking levels? [can select more than one]
      - No
      - Reduced stock volumes [specify 3 main items reduced in volume]
      - Increased stock volumes [specify 3 main items increased in volume]

d. Have you had to change your meeting routine? Yes/ no
   If yes, describe [open-ended]

7. Have you observed any changes in client flow due to the COVID19 outbreak? [select one]
   No
   Decreased client flow overall
   Increased client flow overall
   Both decrease and increase in different client categories

   a. If both decrease and increase,
      i. Which client category(ies) have decreased? [open-ended]
8. Which client category(ies) have increased? [open-ended]

**SECTION III: PROVISION OF COVID19-RELATED SERVICES AND PRODUCTS**

9. Which COVID-related equipment and materials does your pharmacy provide? [indicate number of clients in the last one week]
   None
   Cloth masks [number clients]
   Medical masks [number clients]
   N95/ K95 masks/ respirators [number clients]
   Goggles [number clients]
   Face shields [number clients]
   Gowns/ coveralls [number clients]
   Hand sanitizer [number clients]
   Thermometers [number clients]
   Pulse oximeters [number clients]
   Other, specify type and number of clients
   Specify other COVID19-related equipment and materials that your pharmacy provides

10. Which COVID-related preventive and curative therapies does your pharmacy provide? [indicate number of clients in the last one week]
   None
   Vitamin C [number clients]
   Multivitamins [number clients]
   Traditional/ alternative/ natural remedies, specify [number clients]
   Hydroxychloroquine/ chloroquine [number clients]
   Antibiotics, specify [number clients]
   Antivirals, specify [number clients]
   Immunomodulators, specify [number clients]
   Steroids, specify [number clients]
   Other, specify type [number of clients]
   Specify other preventive and curative therapies provided

   Indicate **number of clients** who bought or requested vitamin C in the last one week
      How many of these vitamin C purchases do you think were COVID-related? [numeric]

   [same format for related subsequent questions]

11. Which COVID-related information and counselling do clients seek from your pharmacy? [indicate number of clients in the last one week]
   None

How to protect themselves from infection [number clients]
Symptoms to look out for if they suspect infection [number clients]
What to do if they develop symptoms or suspect infection [number clients]
Mask use and related problems [number clients]
Counselling for anxiety related to getting infected/ becoming sick with COVID19 [number clients]
Counselling for anxiety about job or business loss [number clients]
Other, specify [number clients]

Specify other type of information or counselling that clients seek: ___________________________

Indicate number of clients in the last one week who asked how to protect themselves from infection

Indicate number of clients in the last one week who asked about symptoms to look out for if they suspect infection

Indicate number of clients in the last one week who asked what to do if they develop symptoms or suspect infection

Indicate number of clients in the last one week who asked about mask use and related problems

Indicate number of clients in the last one week who sought counselling for anxiety related to getting infected or becoming sick with COVID19

Indicate number of clients in the last one week who sought counselling for anxiety about job or business loss

Indicate number of clients in the last one week who sought for other type of information or counselling

12. Select the modes of communication that clients use to seek information/ counselling
    In-person (visit the pharmacy physically)
    Phone calls (including WhatsApp calls etc)
    Texts (including WhatsApp)
    Facebook/ other web-based platforms
    Other, specify

13. What records do you keep regarding COVID-related services and products? [open-ended]
14. Do you prepare any reports regarding COVID-related services and products? Yes/no
    a. To whom do you send the reports?
        Don't send (reports are for internal use)
        Ministry of health (MoH)/ government agency
        Other, specify
    b. What type of information is contained in the reports? [open-ended]

15. What challenges have you had in providing COVID-related services and products? [open-ended]
16. What would you say is your main motivation for supporting delivery of COVID-related services and products? [select one]
        Help people protect themselves/ Help prevent transmission
        Boost the profile of the pharmacy
        Make money
        It's part of my job/ I am required to do it
        Encouragement/ support from professional association (PSK/ KPA/ etc)
        Other, specify
17. How confident do you feel about providing COVID-related services and products? [select one]

Very uncertain/ uncertain/ confident/ very confident

18. What additional support do you feel pharmacies require in provision of COVID-related services and products? [open-ended]
19. If you don't provide COVID-related products or services,
    a. Do you get clients who ask for COVID-related services or products? Yes/no
        i. If yes, what help do you offer them? Referral/ other
            1. If you refer, to where? Another pharmacy/ clinic/ Hospital/ Lab/ other
    b. What COVID-related services or products do clients ask for?
       Personal protective equipment and materials
       Preventive and curative therapies
       Information/ counselling on COVID19
       Other, specify
    c. Select the type of information/ counselling that clients seek, and indicate number of clients in the last one week [select one]
       How to protect themselves from infection [number clients]
       Symptoms to look out for if they suspect infection [number clients]
       What to do if they develop symptoms/ suspect infection [number clients]
       Mask use and related problems
       Counselling for anxiety related to getting infected/ becoming sick with COVID19 [number clients]
       Counselling for anxiety about job/ business loss [number clients]
       Other, specify [number clients]
    d. Select the modes of communication that clients use to seek information, and indicate number of clients in the last one week [select one]
       In-person (visit the pharmacy physically)
       Phone calls (including WhatsApp calls etc)
       Texts (including WhatsApp)
       Facebook/ other web-based platforms
       Other, specify
20. Has any staff member been trained on COVID19?
    a. How many current staff members have been trained on COVID19?
    b. Where did the training take place? On-site/ off-site/ virtual
    c. Who provided the training? MoH/ PSK/ KPA/ Other, specify
21. When you have questions about COVID19, who do you consult? [can select more than one]
    None/ don't consult
    MoH
    Pharmacy in-charge/ manager/ colleague (s)
    Med reps
    Nearby doctor
    Other, specify
22. What is your main source of COVID19 information? [select one]
    TV/ Radio
    Webinar
    Social media
    Google search/ internet
    Scientific journals
    Colleagues/ other pharmacy
    KPA/ PSK communications
    CME/ CPD session
    MoH communications
    Other, specify

**SECTION IV: INFECTION PREVENTION**

23. Which infection prevention measure(s) is your pharmacy using <u>among staff</u>? [can select more than one]
    - None
    - Physical distancing, describe
    - Temperature monitoring
    - Hand hygiene (handwashing, hand sanitizer, etc)
    - Disinfection of surfaces
    - Face masks
    - Goggles
    - Other, specify
24. Which infection control measure(s) is your pharmacy using <u>among clients</u>? [can select more than one]
    - None
    - Physical distancing, describe
    - Temperature monitoring before entering the pharmacy
    - Require clients to be wearing face masks before entering the pharmacy
    - Hand washing
    - Hand sanitizer
    - Use of non-touch payment methods, describe
    - Information materials and messages
    - Other, specify

    Which information materials and messages is your pharmacy providing to clients?
    - Posters/ banners
    - Leaflets/ pamphlets
    - Audio announcements
    - TV screen displays
    - Other, specify

25. Which method do staff members mainly use for cleaning their hands? [select one]
    - Handwashing
    - Hand sanitizer
    - Other, specify

    a. [If handwashing is main] In the last 5 working days,
        i. On how many days was water available for handwashing in your pharmacy? [enter value between 0 and 5]
        ii. On how many days was soap available for handwashing in your pharmacy? [enter value between 0 and 5]
    b. [If hand sanitizer is main] In the last 5 working days,
        i. On how many days was hand sanitizer available in your pharmacy? [enter value between 0 and 5]
26. What type of masks do staff use in your pharmacy? [can select more than one]
    - Cloth masks
    - Surgical/ medical masks
    - N95 or similar
    - Other, specify
27. Have staff members been trained on COVID19 infection prevention? Yes/no
    a. If yes, by who? [can select more than one]
        - MoH
        - Pharmacy in charge/ colleague
        - Other, specify
    b. Where did the training take place? [can select more than one]
        - Off-site, specify venue
        - In-person within the pharmacy

Remotely (virtually) when at the pharmacy
Remotely (virtually) when at home
Other, specify

28. Do you have any written guidelines on COVID19 infection prevention? Yes/no
    a. If yes, describe the guidelines you have access to (e.g. source, title, etc) [open-ended]
29. How adequate do you feel the infection prevention measures in your pharmacy are? [select one]
    Very inadequate/ inadequate/ adequate/ very adequate
30. Has any staff member in your pharmacy ever tested positive for COVID19? [yes/no]
    a. How many staff members have ever tested COVID19 positive? [numeric]
31. State any other comments you have regarding COVID19 infection prevention at your workplace? [open-ended, leave blank if none]

**<u>SECTION V: VIEWS ABOUT CORONAVIRUS 2019 (COV19) TESTING IN PHARMACIES</u>**

32. Do you get clients asking for COV19 testing? Yes/no
    a. In the last one week, how many clients asked for COV19 testing? [numeric]
    b. What help do you offer clients asking for COV19 testing? COV19 tests/ Temperature measurement/ Referral/ other
        i. If referral, where do you refer? [can select more than one]
            Another pharmacy
            Public clinic/ hospital
            Private clinic/ hospital
            Private lab
            Other, specify
33. [If providing COV19 testing] What type of tests do you provide? [can select more than one]
    Antibody rapid tests, specify [unit cost]
    Antigen rapid tests, specify [unit cost]
    Laboratory tests, specify [unit cost]
    Other, specify [unit cost]
34. [If not currently providing] Do you think COV19 testing should be offered in pharmacies? Yes/no
    a. If yes, state the main reason for supporting pharmacy-based COV19 testing [select one]
        There is demand for it/ clients ask for it
        Clients shy away from health facilities
        Clients perceive pharmacies as more confidential compared to other testing facilities
        Pharmacies are open for longer hours
        Pharmacies are easily accessible
        Pharmacies provide speedier (faster) services
        Other, specify
    b. If no, state the main reason for NOT supporting pharmacy-based COV19 testing
        There is no demand for it
        No privacy in pharmacies
        Might be misused
        Other, specify
35. State any other comments you have regarding provision of COV19 testing in retail pharmacies? [open-ended, leave blank if none]

**<u>SECTION VI: OTHER VIEWS AND EXPERIENCES</u>**

36. How prepared do you feel about handling a client who presents to the pharmacy with COVID19 symptoms? [select one]
    Very unprepared/ unprepared/ prepared/ very prepared
37. Which is the main concern you are facing related to personal health and wellbeing? [select one]
    None
    Exposure to infection at work

Lack of transportation to get to work
Being caught up with the curfew
Keeping my family safe
Meeting financial needs for me and my family
Other, specify

38. How much do you feel your personal role as a health worker in the COVID19 response is valued by the community you are serving?
    Not at all valued/ little valued/ highly valued/ very highly valued
39. What other inputs can pharmacies (uniquely) contribute to the national response to the epidemic?? [open-ended]
40. State any other comments you have regarding pharmacy practice during the COVID19 pandemic? [open-ended, leave blank if none]

**<u>Supplementary file B - Simulated client script</u>**

**Call-in simulation**

- Look up the pharmacy on Google Maps and note basic operational details, such as opening hours or COVID19-related client interactions. Take note of any COVID-related messaging.
- Call the customer service number on the Google Maps profile, or if no profile or no number provided, try a simple google search, otherwise contact the study coordinators. They will check if the number in the database is personal or official; if personal further discussion is needed to assess risk of detection.
- **Ask if they have medicines that you can use to protect yourself from getting COVID19, using appropriate language** as agreed during the training. Avoid prompting for information unless it is in the script.
- Make a mental note of **questions asked or advice given**. Encourage the provider to talk by listening actively and saying "don't know" if the provider asks, for example, if you know how to prepare ginger home remedy.
    - If asked why you are asking for the medicines, say that your job requires interaction with customers, some of who don't wear masks
    - If asked where you are located, mention a name of a place near the pharmacy
- If any products are recommended, ask and **write down the names.** You can also ask for the price to keep the conversation natural, but the price data is not being collected
- Ask the service provider **how you can get the medicines**. List down the **options given**, and then if "pick up from the pharmacy" is one of the options, mention that you would rather not visit the pharmacy physically, because you are afraid of getting exposed to COVID19.
- If not told spontaneously,
    - ask what **symptoms you should look out for** if you suspect COVID19 infection? If asked what symptoms you have, emphasize you don't have symptoms but just want to be ready.
    - ask **what you should do if you suspect you have COVID19 infection**?
- Thank the service provider and mention that you will get back to them to arrange how to get the medicines.

**Completing the debrief questionnaire**

- The debrief questionnaire should ideally be completed within 30 minutes of the simulation, while memory is still fresh. In any case, do not call another pharmacy before the previous debrief interview has been done.
- It is **very important to say honestly how the call went**, even if you feel like it didn't go very well. The **study is assessing the pharmacy and not you**, the simulated client.
- Try to capture the responses to open-ended questions close as possible to what the provider says (verbatim)
- If there is follow-up interaction, you can leave the questionnaire "unsubmitted" and finish later. Open a fresh page if you need to do another pharmacy
- If further information comes in after you have submitted, relay to the study coordinators who will then edit the record from the back end.

**Supplementary file C - debrief questionnaire**

Basic data

- Pharmacy identifier
- Simulated client identifier
- Simulation type: in-person/ call-in

Call-in simulation

- What time did you call the pharmacy? [24-hour format]
- What was the gender of the person who served you? Male/ female/ could not tell
- Were any medicines recommended? Yes/no
- If yes, which medicines were recommended for prevention? [Indicate price for each]
    - None
    - Vitamin C
    - Zinc
    - Multivitamins
    - Azithromycin,
    - Other antibiotics (Augmentin, Amoxil, ceftriaxone, etc)
    - Herbal medicine, specify
    - Home remedies, specify
    - Other, specify
- What options were given on how you can get the recommended medicines?
    - None
    - Come to the pharmacy and pick
    - Send someone to the pharmacy to pick
    - The pharmacy to deliver to client's location
    - From nearest chemist/ health facility
    - Other, specify
- If no medicines recommended, what was recommended?
    - None
    - Mask use
    - Hand hygiene (washing, sanitizer)
    - Other, specify
- Were you asked why you are asking for the medicines? (yes/no)
- Before prompting, were you told the symptoms to look out for if you suspect COVID19 infection? Yes/no
    - If yes, describe [open-ended]
- After prompting, what symptoms were you told to look out for if you suspect COVID19 infection? [open-ended]
- Before prompting, were you told what to do if you suspect you have COVID19 infection? Yes/no
    - If yes, what was recommended if you suspect COVID19 infection?
        - Medications
        - Visit nearest health facility/ go see a doctor
        - Check for information online/ WHO website
        - Get tested for COVID19
        - Get checked for oxygen level
        - Get checked for body temperature
        - Self-quarantine/ isolate at home
        - other, specify
    - If medications, what medications were recommended for treatment?
        - Steriods (dexamethasone, betamethasone, prednisolone, etc)
        - Azithromycin,

        - Other antibiotics (Augmentin, Amoxil, ceftriaxone, etc)
        - Painkillers (paracetamol, brufen, etc)
        - Herbal medicine, specify
        - Home remedies, specify
        - Other, specify
- After prompting, what were you told to do if you suspect you have COVID19 infection? Medications
        - Nothing
        - Visit nearest health facility/ go see a doctor
        - Check for information online/ WHO website
        - Get tested for COVID19
        - Get checked for oxygen level
        - Get checked for body temperature
        - Self-quarantine/ isolate at home
        - other, specify
    - If medications, what medications were recommended for treatment?
        - Steriods (dexamethasone, betamethasone, prednisolone, etc)
        - Azithromycin,
        - Other antibiotics (Augmentin, Amoxil, ceftriaxone, etc)
        - Painkillers (paracetamol, brufen, etc)
        - Herbal medicine, specify
        - Home remedies
        - Other, specify
- What other advice was given?
        - None
        - Wear a mask
        - Sanitize/ wash hands
        - Eat fruits
        - Other, specify
- What other questions were you asked? [open-ended]
- About how long was the call? [mins]
- Was there further interaction after the call? Yes/no
    - If yes, describe
        - SMS
        - Whatsapp
        - Follow-up call
        - Other, specify

<u>General observations</u>

- How friendly (approachable) was the person (s) who served you?
  1 – Very unfriendly
  2 – Unfriendly
  3 – Friendly
  4 – Very friendly
- How enthusiastic was the provider about providing the service?
  1 – Very unenthusiastic
  2 – Unenthusiastic
  3 – Enthusiastic
  4 – Very enthusiastic
- Did the provider say or do anything that you found particularly helpful or nice? Yes/no
  If yes, describe [free text]
- Did the provider say or do anything that you found particularly NOT helpful or NOT nice? Yes/no
  If yes, describe [free text]

- Do you have any additional observations about the interaction? Yes/no
    If yes, describe [free text]

**Supplementary file D - In-depth Interview Topic Guide:**

**Assessing the response to the COVID-19 epidemic by private retail pharmacies in Kenya.**

**Notes to interviewer:**

1. As first option, use Teams platform with video on. If internet is slow, try Teams without video and if that still doesn't work then use plain old telephone (POT).
2. If using POT, put the phone on speaker and record on a standard audio recorder placed next to the phone, and if using Teams, record directly through the application.
3. When setting up the interview, ask about internet access, preference of platform, etc. Inform the participant that they will need to be in a quiet place for about 30-60 minutes and that earphones are preferred to speaker mode.
4. Information required from the questionnaire:
    a. Preparedness level in handling COVID suspect case
    b. Products provided
    c. Information and counseling provided
    d. Whether they get clients who ask for testing
    e. Whether they think testing can be offered in a pharmacy

*We would like to understand how the pharmacy sector is responding to the COVID19 epidemic, including: initial impact, experiences providing services during the epidemic, and views about pharmacy-based COVID19 testing. Towards the end of the interview I will also ask you a few questions about your experience being interviewed remotely compared to face-to-face.*

**Interviewee profile**

1. Briefly tell me about your pharmacy (setting, staff, clients)?
    - What is your role in the pharmacy?

**Initial response and impact**

2. When did you first hear about COVID19? From who?
    - What did you think and feel about it at that time?

3. What was the initial response at your pharmacy?

4. What was the effect of the epidemic on pharmacy operations?
    - Client flow, sales, working hours, staff, meeting routine, etc

5. What has been your role in responding to the epidemic?
    - You said your preparedness in handling a COVID-19 case was: _________________. Please explain further
    - What aspects of your education and previous experience helped you in responding to the outbreak?

**COVID-related products and services**

6. In the questionnaire, you mentioned a number of products that your pharmacy is providing in relation to the epidemic. ________________________________________________________________________________
   Which of these products were introduced specifically because of the epidemic?
    - You also mentioned information and counseling that clients are seeking or you are providing? ____________________________________________________________________________
    - How is this done?
    - How does your pharmacy handle suspected COVID19 cases?

7. Have you been involved in any COVID training or peer learning sessions, either as a participant or a trainer?

- If yes, Where? When? Who else was included?
    - Was it stand alone training on this topic or part of a wider training?
    - Do you feel the training was adequate?
    - Who was supporting the training (financially and technically)?
- If no, how did you learn how to provide COVID services and products?

8. What do you think is needed to improve the delivery of COVID-related services in the retail pharmacy sector?
    - What support (material or financial) can be given by the government or any other external party? (e.g., drugs and other supplies/ toolkits/ job-aids/ digital systems)
    - Do you report information on COVID-related services to the government or any other external party?

**<u>Pharmacy-based COVID19 testing</u>**

9. If they get clients who ask for COVID19 testing: YES NO (*circle one*)
    - What type of clients ask for testing? (reasons for wanting to test, etc)
    - How do you help clients asking for COVID19 testing?
    - Why do you think they prefer pharmacies rather than health facilities or other testing sites?

10. You mentioned that COVID19 testing SHOULD/ SHOULD NOT *(circle one)* be provided in pharmacies? Explain.
    - What would be needed to ensure successful implementation of COV19 testing in the retail pharmacy sector?
    - What type of tests would be most suited for pharmacy-based COVID19 testing?

**<u>Other potential inputs into the response</u>**

11. What other inputs can pharmacies (uniquely) contribute to the national response to the epidemic?
    - How would this be done?

**Conclusion of main interview**: We have come to the end of the specific questions I had. Do you have any further comments?

**<u>Experience doing the interview remotely</u>**

*Now I would like to ask you a few questions about your experience being interviewed remotely compared to face-to-face.*

12. How would you describe your experience doing the interview remotely, compared to face-to-face?
    - Have you done a face-to-face interview before? Describe.
    - Advantages and challenges
    - Would you have preferred a face-to-face interview? Explain.

13. What did you like or not like about the Teams platform?
    - Do you have previous experience with digital meeting applications? (Teams, Zoom, etc)
    - Would you have preferred another platform?

**Conclusion**: Thank you very much for your time. If it is ok with you, I may contact you again for any clarifications or to hear about additional experiences and views on the topic. We will share a report once data analysis is complete.

**Supplementary file E - Checklist – Consolidated criteria for reporting qualitative studies (COREQ)**[i]

**Manuscript:** Response to the coronavirus disease 2019 (COVID-19) pandemic at private retail pharmacies in Kenya: a mixed methods study

| Item number | Guide questions/description | Reported on Page # |
|---|---|---|
| **Domain 1: Research team and reflexivity** | | |
| *Personal Characteristics* | | |
| 1. Inter viewer/facilitator | Which author/s conducted the interview or focus group? | 4 |
| 2. Credentials | What were the researcher's credentials? E.g. PhD, MD | The names and affiliations are included on title page |
| 3. Occupation | What was their occupation at the time of the study? | The names and affiliations are included on title page |
| 4. Gender | Was the researcher male or female? | One female, one male |
| 5. Experience and training | What experience or training did the researcher have? | The names and affiliations are included on title page |
| *Relationship with participants* | | |
| 6. Relationship established | Was a relationship established prior to study commencement? | 4- Participants had been included in a previous study |
| 7. Participant knowledge of the interviewer | What did the participants know about the researcher? e.g. personal goals, reasons for doing the research | 5 – participants signed a consent form that explained the research goals |
| 8. Interviewer characteristics | What characteristics were reported about the inter viewer/facilitator? e.g. Bias, assumptions, reasons and interests in the research topic | No bias to report |
| **Domain 2: study design** | | |
| *Theoretical framework* | | |
| 9. Methodological orientation and Theory | What methodological orientation was stated to underpin the study? e.g. grounded theory, discourse analysis, ethnography, phenomenology, content analysis | 5- thematic analysis |
| *Participant selection* | | |
| 10. Sampling | How were participants selected? e.g. purposive, convenience, consecutive, snowball | 4- purposive |
| 11. Method of approach | How were participants approached? e.g. face-to-face, telephone, mail, email | 4- telephone |
| 12. Sample size | How many participants were in the study? | 5 – 18 participants |
| 13. Non-participation | How many people refused to participate or dropped out? Reasons? | Participatin rate was 56% of 32 providers invited; 7 of those invited did not respond after three reminders, 5 said they were not interested, and 2 had already left the pharmacy |

| Item number | Guide questions/description | Reported on Page # |
|---|---|---|
| | | *Not included due to word limit* |
| *Setting* | | |
| 14. Setting of data collection | Where was the data collected? e.g. home, clinic, workplace | 4-phone |
| 15. Presence of non-participants | Was anyone else present besides the participants and researchers? | No.<br><br>*Not included due to word limit* |
| 16. Description of sample | What are the important characteristics of the sample? e.g. demographic data, date | 5 – supp tab 2 |
| *Data collection* | | |
| 17. Interview guide | Were questions, prompts, guides provided by the authors? Was it pilot tested? | 4 – supp file D<br><br>The interview guide was updated after the first few interviews based on responses from participants and how effectively these addressed research objectives.<br><br>*Not included due to word limit* |
| 18. Repeat interviews | Were repeat interviews carried out? If yes, how many? | No repeat interviews were done.<br><br>*Not included due to word limit* |
| 19. Audio/visual recording | Did the research use audio or visual recording to collect the data? | 5 – data management and analysis |
| 20. Field notes | Were field notes made during and/or after the interview or focus group? | Field notes made after the interviews<br><br>*Not included due to word limit* |
| 21. Duration | What was the duration of the interviews or focus group? | Interviews lasted a median [range] of 35 [20-92] minutes<br><br>*Not included due to word limit* |
| 22. Data saturation | Was data saturation discussed? | To expedite data collection, all 18 interviews were completed before starting analysis or checking for saturation<br><br>*Not included due to word limit* |
| 23. Transcripts returned | Were transcripts returned to participants for comment and/or correction? | Transcripts were not returned to participants for comment and/or correction |

| Item number | Guide questions/description | Reported on Page # |
|---|---|---|
| | | *Not included due to word limit* |
| **Domain 3: analysis and findings** | | |
| *Data analysis* | | |
| 24. Number of data coders | How many data coders coded the data? | 5 – data management and analysis |
| 25. Description of the coding tree | Did authors provide a description of the coding tree? | No.<br><br>*Not included due to word limit* |
| 26. Derivation of themes | Were themes identified in advance or derived from the data? | Analysis followed a thematic approach with initial categories based on the interview guide and emergent themes integrated<br><br>*Not included due to word limit* |
| 27. Software | What software, if applicable, was used to manage the data? | 5- NVivo 12 |
| 28. Participant checking | Did participants provide feedback on the findings? | Participants were not asked for feedback on the findings.<br><br>*Not included due to word limit* |
| *Reporting* | | |
| 29. Quotations presented | Were participant quotations presented to illustrate the themes/findings? Was each quotation identified? e.g. participant number | Yes. Results section |
| 30. Data and findings consistent | Was there consistency between the data presented and the findings? | Yes. |
| 31. Clarity of major themes | Were major themes clearly presented in the findings? | Yes. Results section |
| 32. Clarity of minor themes | Is there a description of diverse cases or discussion of minor themes? | Yes. Results section |

[i] Developed from:
Tong A, Sainsbury P, Craig J. Consolidated criteria for reporting qualitative research (COREQ): a 32-item checklist for interviews and focus groups. *International Journal for Quality in Health Care*. 2007. Volume 19, Number 6: pp. 349 – 357
